# Focusing on Patient-outcome performance measures of Active and Passive Implants – A Systematic Review

**DOI:** 10.1101/2022.05.30.22275158

**Authors:** Jana Stucke, Elinor Tzvi-Minker, Andreas Keck

## Abstract

**Motivation:** Active implantable electronic medical devices are used in different fields of medicine, in particular cardiology and neurology. Several papers have been published over the years comparing the technical performance of implants between leading manufactures. However, no such comparison has been done with respect to “Patient-Reported Outcomes” (PROs) for most implant types, despite its importance in evaluating the quality of a medical device. With the recent update of the European Union’s (EU) regulation on public procurement towards value-based medicine, it has become beneficial for manufacturers to focus more on PROs to differentiate their products in order to create a marketing leverage. Most importantly, investigating PROs can assist shared decision-making, support pharmaceutical labelling claims and influence healthcare policy and practice. Due to this the review aims to showcase the lack of PRO comparisons between implant manufacturers across different medical fields and its impact on patients and surgeons.

**Methods:** A literature search was conducted for active and passive implant performance comparisons in the area of implantable cardioverter-defibrillator (ICDs), deep brain stimulation (DBS), cochlear implants (CIs) and intraocular lenses (IOLs). The search focused on the availability of manufacturer comparisons with regards to PROs. A total of 640 papers from 2000 until 2022 were screened in detail for the search term “patient reported outcomes” for the different implant types. Next, we analyzed the results by tagging papers based on the specific topics they investigated in their study to enable a cross-comparison. We noted whether the implant manufacturers were mentioned as well as whether a manufacturer comparison was done. Studies were also evaluated based on the number of patients included.

**Results:** A total of 38 papers were identified for ICDs, 31 for DBS, 68 for IOLs and 34 for CIs. 50% of the papers for IOL focused on PROs and 22% and 14% for CIs and DBS, whilst mentioning the manufacturers. No papers dealing with PROs could be identified for ICDs. Manufacturer comparison was not attempted by any of the reviewed papers, despite implants such as CIs and DBS having quite a significant impact on the quality of life.

**Conclusions:** There is an urgent need for clinical studies which focus on PRO comparisons between implants of different manufacturers, to not only provide physicians and patients with critical information that informs their decision prior to implantation, but also to increase the competition between manufacturers, thus, fostering innovation. The former would allow physicians to recommend the most suitable implant for the patient. In addition, this will drive manufacturers towards PRO focused improvements.

## Introduction

The use of implantable active and non-active medical devices has increased dramatically in the last few decades. Due to advancements in technology and especially Artificial Intelligence (AI), current state-of-the-art devices can treat conditions that have not been treatable up until now before. These advancements have led the highly competitive medical manufacturing industry to differentiate their products by demonstrating product value beyond the traditional measures of clinical safety and efficacy (Curran, 2018; Squitieri et al., 2017), through patient-reported-outcomes (PROs).

A PRO is defined as a report of the status of the patient’s health condition, by the patient itself or their caregiver or surrogate responder, without prior interpretation (Eton et al., 2014; U.S. Food and Drug Administration, 2009; European Medicines Agency, 2019). It classifies a range of different, patient-related concepts, including personal reports of health status, symptoms, and health-related quality of life (QoL, Calvert et al., 2013). PROs allow for shared decision making between practitioner and patient, which makes them a valuable marketing tool for medical device manufacturers.

One sector within the medical device industry, which may profit most from the use of PROs are medical implants. Passive implants have been used since the mid 1950’s in the form of breast implants and quickly been followed by active implants, such as pacemakers in 1958. Nowadays numerous passive and active implants from head to toe exist, from dental to hip implants as well as cardiac defibrillators (ICDs) and Cochlear implants (CIs). It is predicted that the global active implantable medical device market will increase its CAGR by 7.7% (Market Data Forecast) by 2026. With personalised medicine gaining more importance due to the rise of AI, medicine in general has become more customer centric. Patient Reported Outcomes (PROs), which assess the impact of a medical treatment or intervention (Rothman et al., 2007), becomes a decision factor for surgeons and patients alike. This is particularly true for implants as they require an intervention and, depending on the implant, most often cannot be easily replaced.

For years, studies have been conducted comparing the technical features of different active and passive implant manufacturers in different medical fields (Saad et al., 2002; Semmler et al., 2021; Mens 2007; Plontke et al. 2018; Bon et al, 1997; Nagy et al., 2011). In particular, transcatheter pacemakers and ICDs have been investigated, and the focus of these studies was mainly on lead malfunctions and battery longevity (Ipp, 2021; Hauser et al, 2021, De Meester and Gilles, 2001; Narayanan et al., 2021, Pokorney et al., 2014). However, studies performing outcome-based manufacturer comparisons are rare. This is true for most implant types apart from breast implants, which are commonly used for plastic surgery. Here, a vast number of publications comparing manufacturers can be found (Niechajev et al., 2007; Yoon and Chang, 2020; Henderson et al., 2015). However, active implants may be an intervention with a more drastic impact on a patients’ QoL. The implantation of CIs, for example, requires the insertion of an electrode array into the cochlea, whilst deep brain stimulation (DBS) technology requires an accurate placement of the leads in the brain and a personalization of the stimulation parameters according to the severity of the neurological symptoms. Thus, it is of critical importance, especially in these particular areas of active implants to: (1) report systematically on PROs and (2) perform objective manufacturer comparisons that could guide patients and surgeons alike in their decision-making process.

In this review, we scanned the literature for studies of both active and passive implants that report patient outcomes. In the area of active implants, we focused on implanted cardiac defibrillators (ICDs), cochlear implants (CIs) and deep brain stimulation (DBS), whereas for passive implants we focused on intraocular lenses (IOLs). The latter was chosen due to its similarity to CIs in affecting sensory perception, and thus PRO. Note that these are exemplary passive and active implants, and there are many more medical implants that could be investigated in future work, such as artificial knees and hips as well as leadless and transvenous pacemakers. The purpose of this review is to demonstrate the availability of manufacturer information in different active and passive implants and to stress the importance of performing comparisons of PROs in the different medical fields.

## Methods

### Search Strategy

A structured literature search was conducted using both google scholar and PubMed. The search strategy comprised searching for free-text terms “patient reported outcomes”, combined with the different implanted devices: “ICD”, “Cochlear Implant”, “intraocular lenses” and “deep brain stimulation”. An initial search found that PubMed showed less relevant hits than Google Scholar and the hits found in the former matched those found on Google Scholar. To this end, two independent reviewers (ETM, JS) scanned the first 150 most relevant (according to Google Scholar relevance algorithm) hits. We selected all papers published after 2000 in peer-reviewed journals. All reviews and commentary articles were discarded.

The literature search was last updated 22nd February 2022. The search targeted published research in scholarly journals, conference proceedings and workshop reports that assesses active or passive implants on a technical or outcome-based basis.

### Selection Process

Titles, abstracts, and full articles were subsequently screened by reviewer 1 [JS] applying the inclusion and exclusion criteria (Figure 1). Publications meeting the inclusive criteria, were reviewed a second time by an additional reviewer [ETM]. Figure 1 illustrates the process.

**Figure 1.**
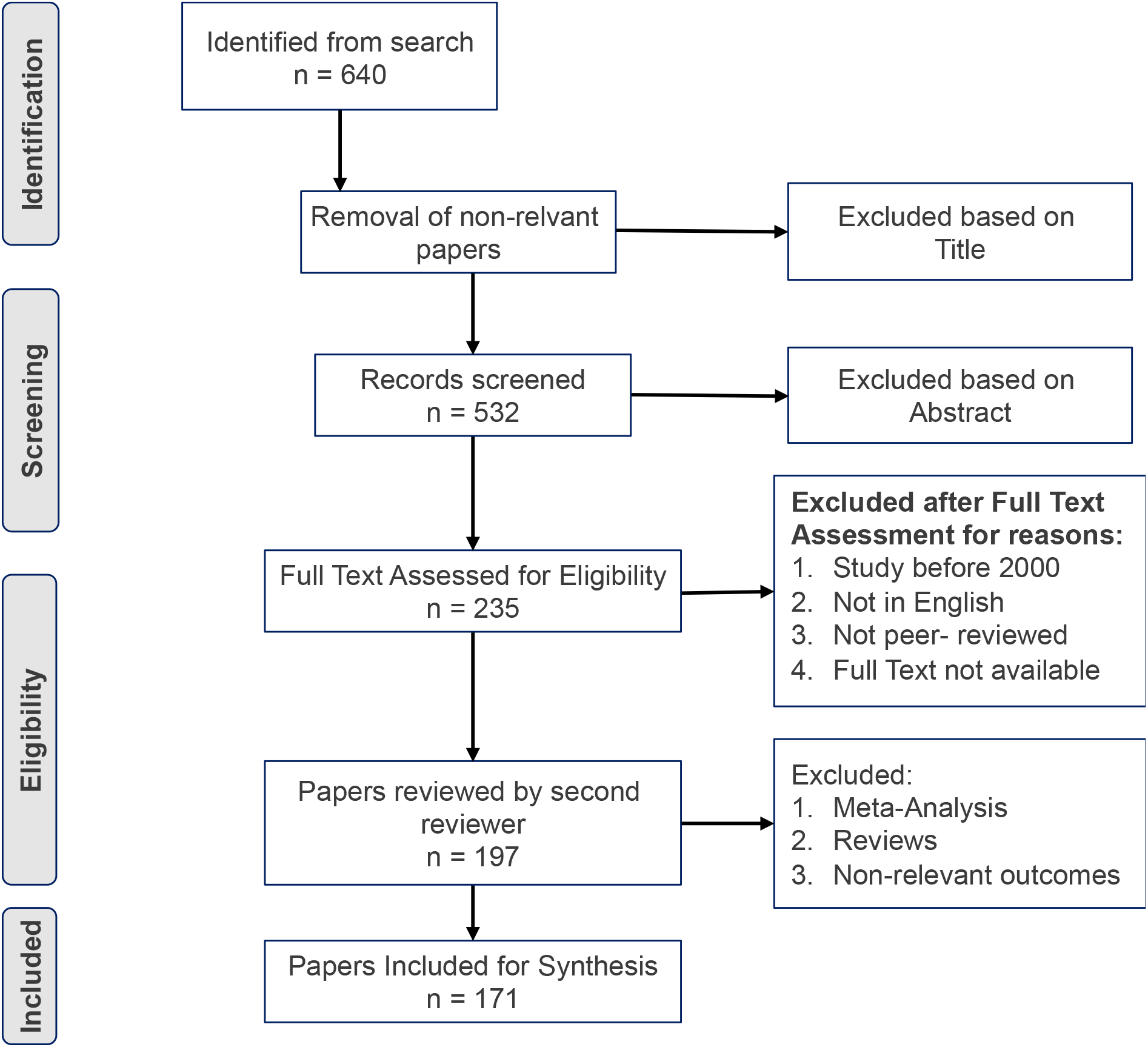
Literature Inclusion Flowchart.

### Data Extraction and analysis

Data was extracted from the included papers in a matrix. The data extraction was mainly done by reviewer 1 [JS] and later re-examined by reviewer 2 [ETM]. The extracted data was categorised and summarised in a matrix and later exported into tables and graphs. The data matrix is provided for a convenient workflow in Microsoft Excel (version 16.16.5) and can be found in the Appendix.

## Results

The following section presents a summary of the extracted data from 171 included studies investigating PROs of active and passive implants for the areas of neurology, cardiology, and otolaryngology. Specifically, we focused on the following devices: implanted cardiac defibrillators (ICDs), cochlear implants (CIs) and deep brain stimulation (DBS), whereas for passive implants we focused on intraocular lenses (IOLs).

### Bibliographic Overview

Table 2 shows the number of total publications considered after the reviewing process. Here, we included publications from peer-reviewed journals only. Conference proceedings, workshop publications or case studies were not considered. The Journal of American College for Cardiology (n = 9), Otology & Neurotology (n = 4), Journal of Cataract & Refractive Surgery (n = 22) and Brain (n = 4) represented the journals with most included publications for ICDs, CIs, IOLs and DBS respectively.

**Table 1.** Bibliographic overview of the included studies

| Implant Type | Number of papers before filtering | Number of papers after filtering |
| --- | --- | --- |
| ICD | 47 | 38 |
| DBS | 40 | 31 |
| IOL | 89 | 68 |
| CI | 59 | 34 |

**Table 2.** Maximum, Minimum and Average Number of Patients used in the Literature for ICDs, CIs, DBS and IOLs

| Implant Type | Maximum Number of Patients | Minimum Number of Patients | Average Number of Patients |
| --- | --- | --- | --- |
| ICD | 38,912 | 12 | 3,142 |
| CI | 2,735 | 19 | 180 |
| DBS | 5,311 | 3 | 237 |
| IOL | 710 | 6 | 117 |

The included papers were published during the following years for ICDs: 2012 (n = 9), 2015 (n = 4) and 2005 (n = 4); IOLs: 2017 (n = 13), 2021 (n = 12) and 2019 (n = 10); DBS: 2010 (n = 6), 2006 (n = 4) and 2012 (n = 4) and CIs; 2021 (n = 16), 2020 (n = 16) and 2019 (n = 3). This shows that the general interest in PROs is clearly growing for CI and IOLs when compared to DBS and ICDs.

### Sample Sizes

The total number of subjects in the included studies varied with implant type with the maximum, minimum and average patient number for each implant being depicted in Table 2. Note, that for ICDs eight studies were retrospective, analysing very large patient databases ranging from 1,820 to 38,912 patients. Large retrospective studies were also performed in the other fields. For DBS, one study had over 5,000 participants, and for CI two studies had more than 1,500 participants. For IOLs, only three studies had over 500 participants. This results in a high average of participants for ICDs. The other implant types had on average between 117 and 237 patients, which can be considered quite high and reflecting the quality of the paper considered in this review. Only DBS and IOL included cohorts of less than 12 subjects which may be due to a specific focus on aetiology.

### Summary of findings in the Literature

A distribution of the papers according to outcome, as tagged in Table 3, is shown in Figure 2 to Figure 5. This was done for ease of comparison across implant types. Here, papers were tagged based on their overarching topic for ease of cross-comparison in the different medical fields.

**Table 3.** Legend of the taxonomy used for study tagging

| Overarching Term | Tagging of papers based on |
| --- | --- |
| Safety | Safety of the implant as defined by the authors |
| QoL | “Quality of Life” directly mentioned in the text or<br>PRO assessed as QoL measure |
| Complications | Side effects as well as mortality |
| Psychological | Anxiety, Fear, Depression |
| Effectiveness | Effectiveness of the implant to treat the patient |
| Cognitive Improvement | Any type of cognitive ability that improved following implantation |
| Rehabilitation | Studies focusing on best practices for post-surgery rehabilitation |
| Performance | Patients' improvement in test scores |
| Satisfaction | Patient satisfaction with the outcome |
| PRO | A clear statement of Patient Reported Outcomes in the text |
| Comparison hearing ability criteria | Only used for CIs. Evaluation of the tests to judge whether an implantation was beneficial |

**Figure 2.**
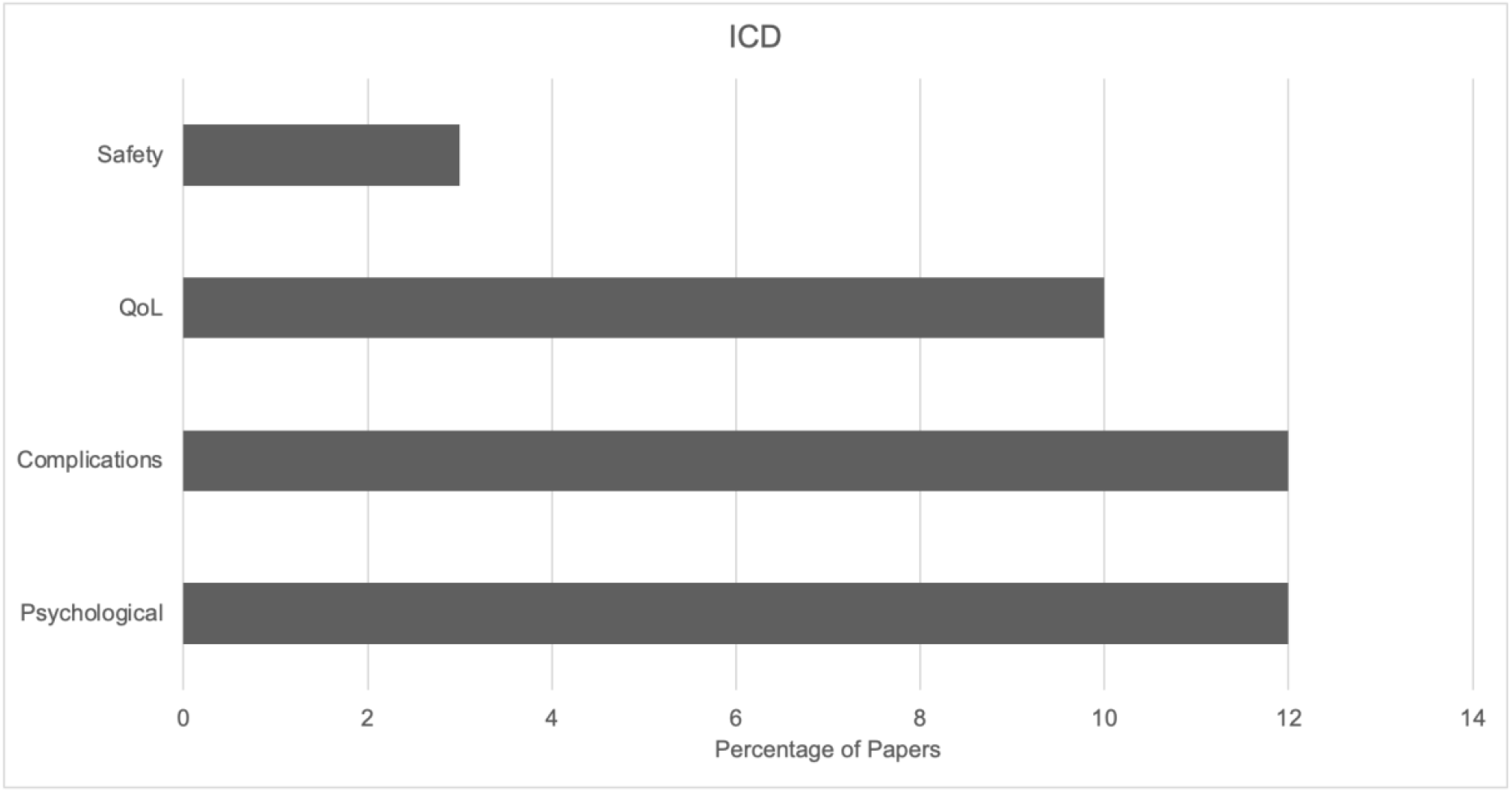
Occurrence of the created taxonomy within the screened ICD studies.

**Figure 3.**
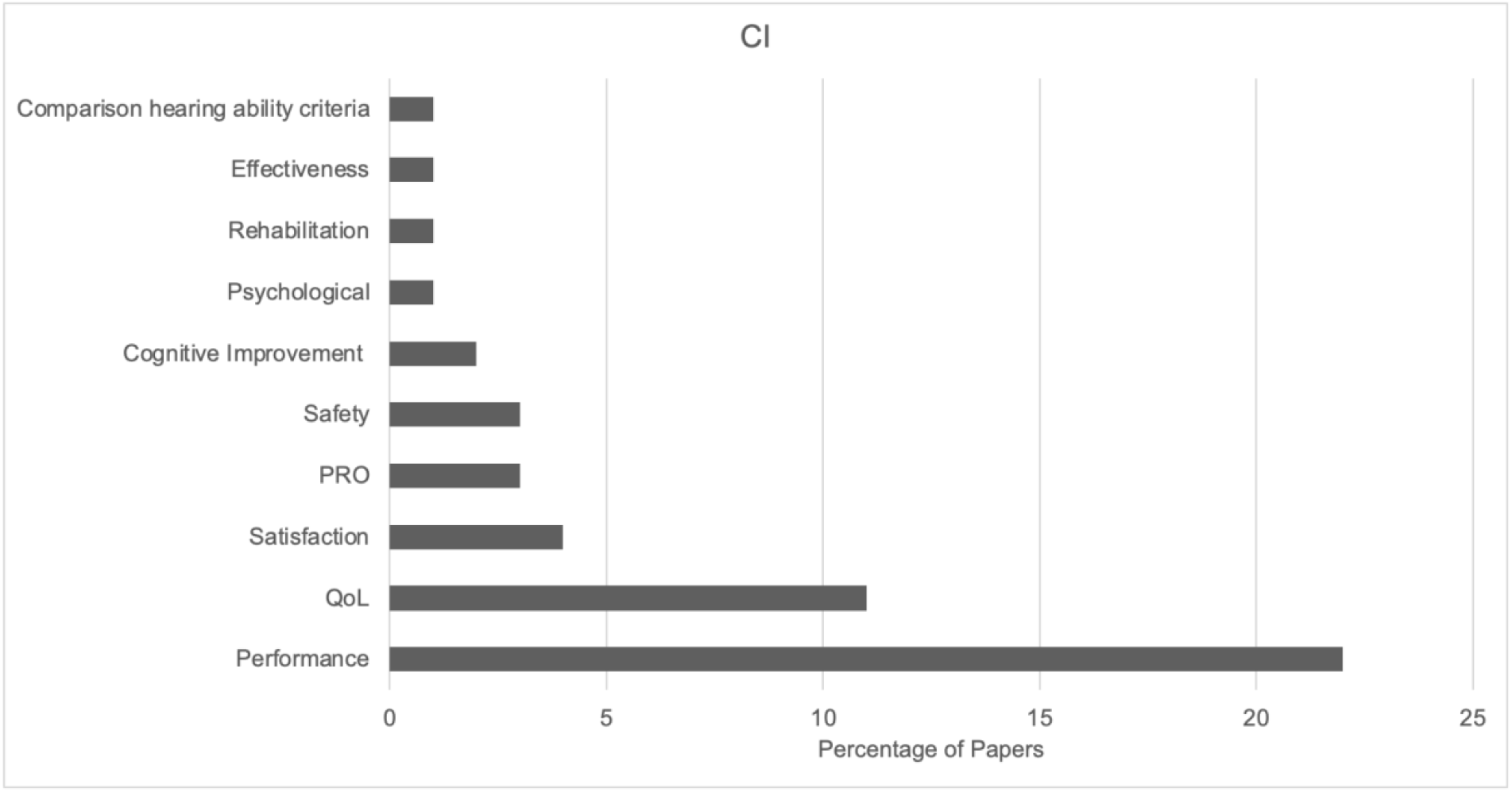
Occurrence of the created taxonomy within the screened CI studies.

**Figure 4.**
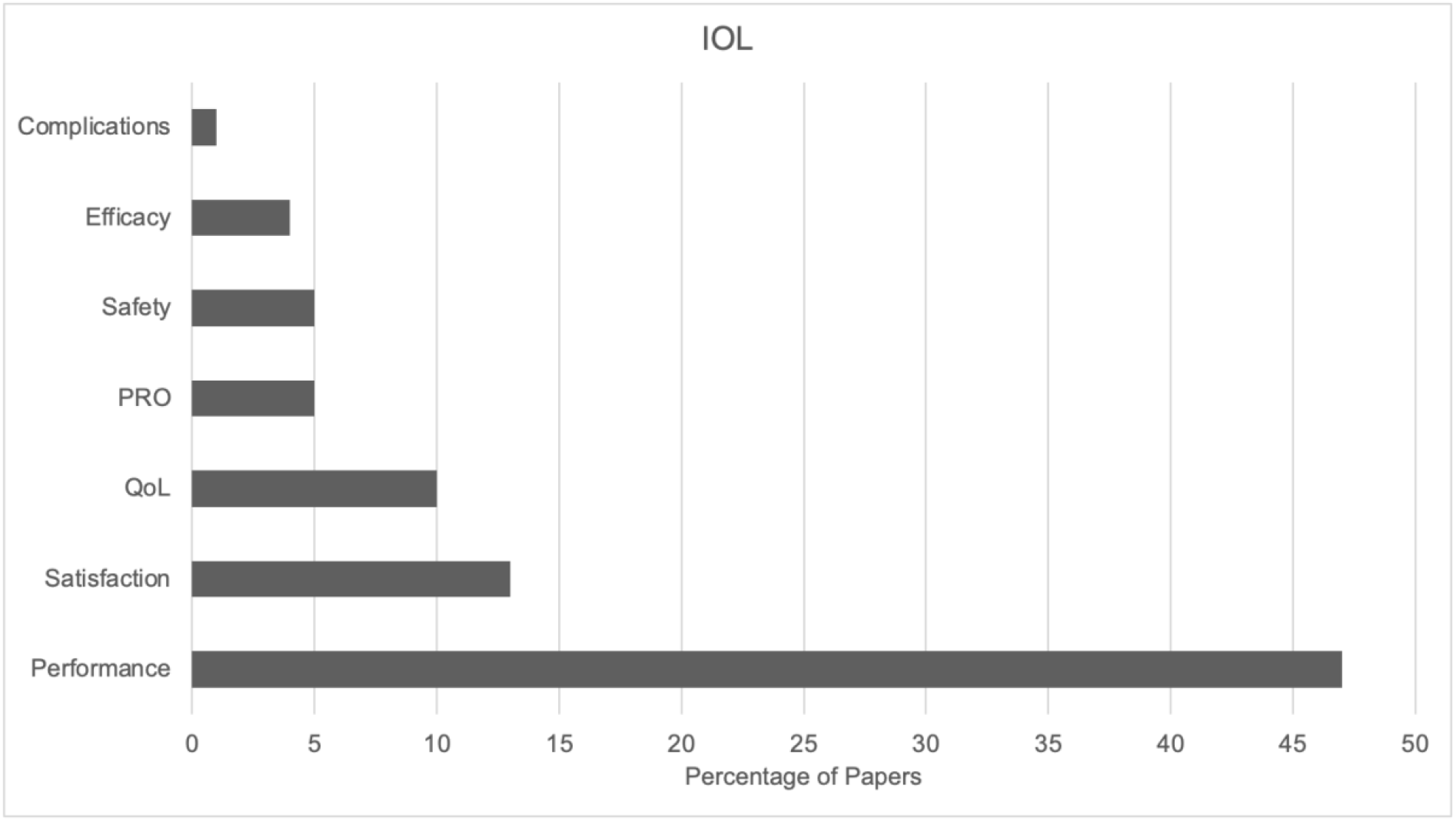
Occurrence of the created taxonomy within the screened IOL studies.

**Figure 5.**
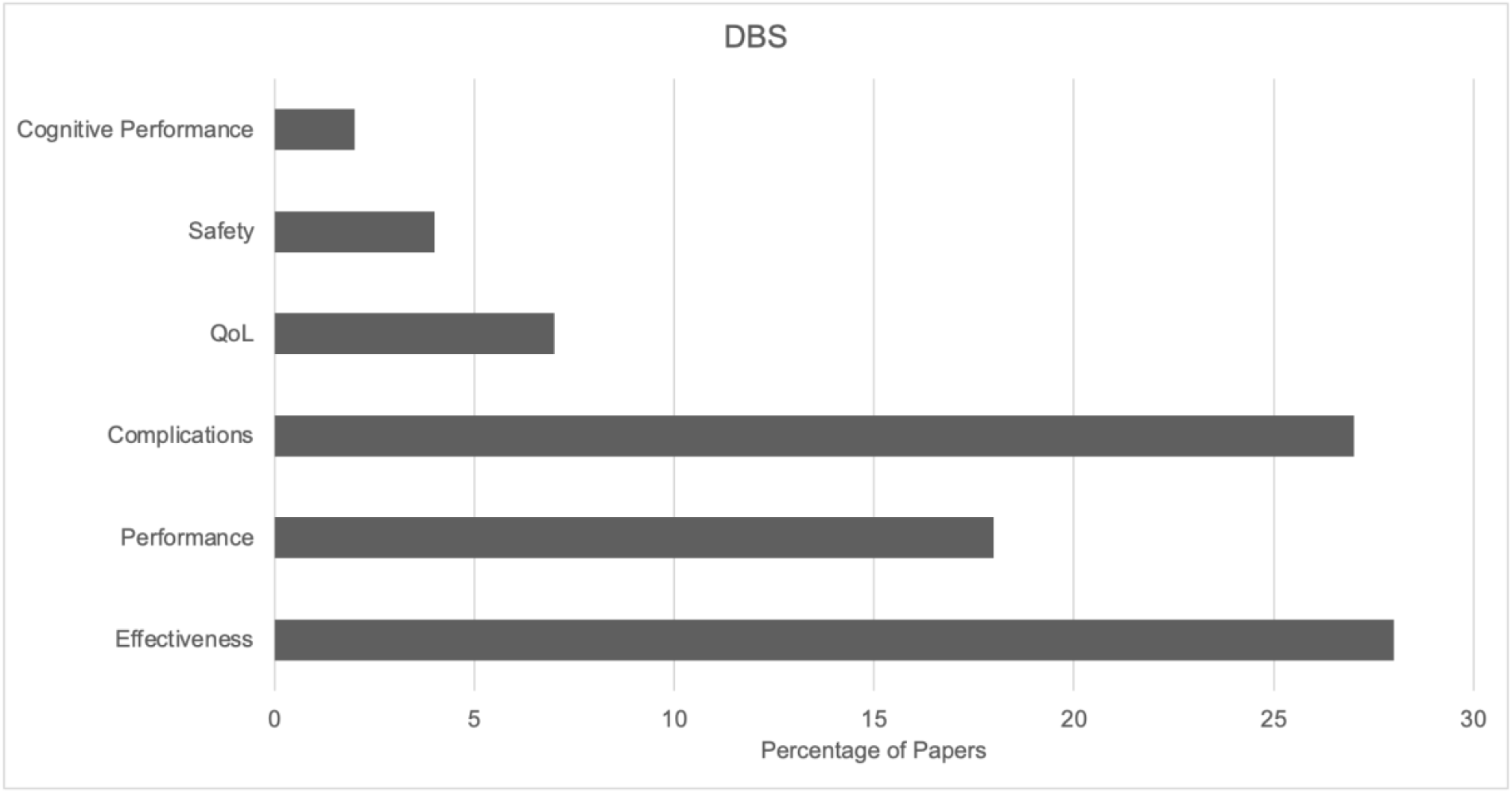
Occurrence of the created taxonomy within the screened DBS studies.

The results show that most papers evaluating PROs for ICDs deal with the psychological effect on the patient, as well as the associated complications of the implantation. We can therefore conclude that these outcomes serve as most significant in this medical field. Several papers (n = 10) also investigated QoL as the main outcome, which is tightly knit to psychological outcomes.

For CIs the distribution of topics is less balanced than for ICDs. Here, there is a heavy focus on the patient’s performance, in particular speech and hearing perception scores. These scores reflect how well the patient functions with the implant and serve as a comparison to the pre-operative state, thus, can be seen as closely related to QoL. PROs and satisfaction played only a minor role in the research.

Figure 4 shows the distribution of outcomes investigated for IOLs. As for CIs, most studies deal with performance, with satisfaction and QoL lagging much behind. This distribution is very different to the more balanced distribution of outcomes for ICDs.

The outcome distribution for DBS is depicted in Figure 5. It can be seen that over 35% of the papers are addressing the effectiveness of DBS to treat different conditions. Performance, which means in this case, how well the patients improved their symptoms in each indication (Parkinson’s disease, Dystonia, Essential tremor, Obsessive compulsive disorder and depression) following implantation, and complications are often mentioned alongside effectiveness and are forming the second and third most mentioned topics in the DBS studies reviewed here. Only very few studies evaluated QoL directly, though effectiveness of the therapy is also an indirect indication of QoL.

Figure 6 shows a cross-comparison of outcomes for the different implant types. We found that the focus of ICD papers lies heavily on the psychological impact on the patient and complication rate, whereas CI and IOL papers are mainly addressing performance criteria. DBS studies are mostly concerned with the effectiveness of treatment compared to other forms of therapies. Interestingly, there are relatively more papers focusing on QoL for ICDs than for CIs, DBS and IOLs, despite the latter directly impacting perception (visual, hearing, motor). It is also surprising that double the amount of papers on IOLs focus on patient satisfaction than for CIs. Here, a more balanced result would have been expected, in particular, as CI surgery affects hearing perception and similar to IOLs directly affects sensation. The same is true for DBS, which is used for alleviating neurological symptoms in Parkinson’s disease and depression. There as well, one would expect more studies investigating patient satisfaction. Generally, as these disorders have a strong effect on quality of life, more papers on the assessment of QoL would have been expected.

**Figure 6.**
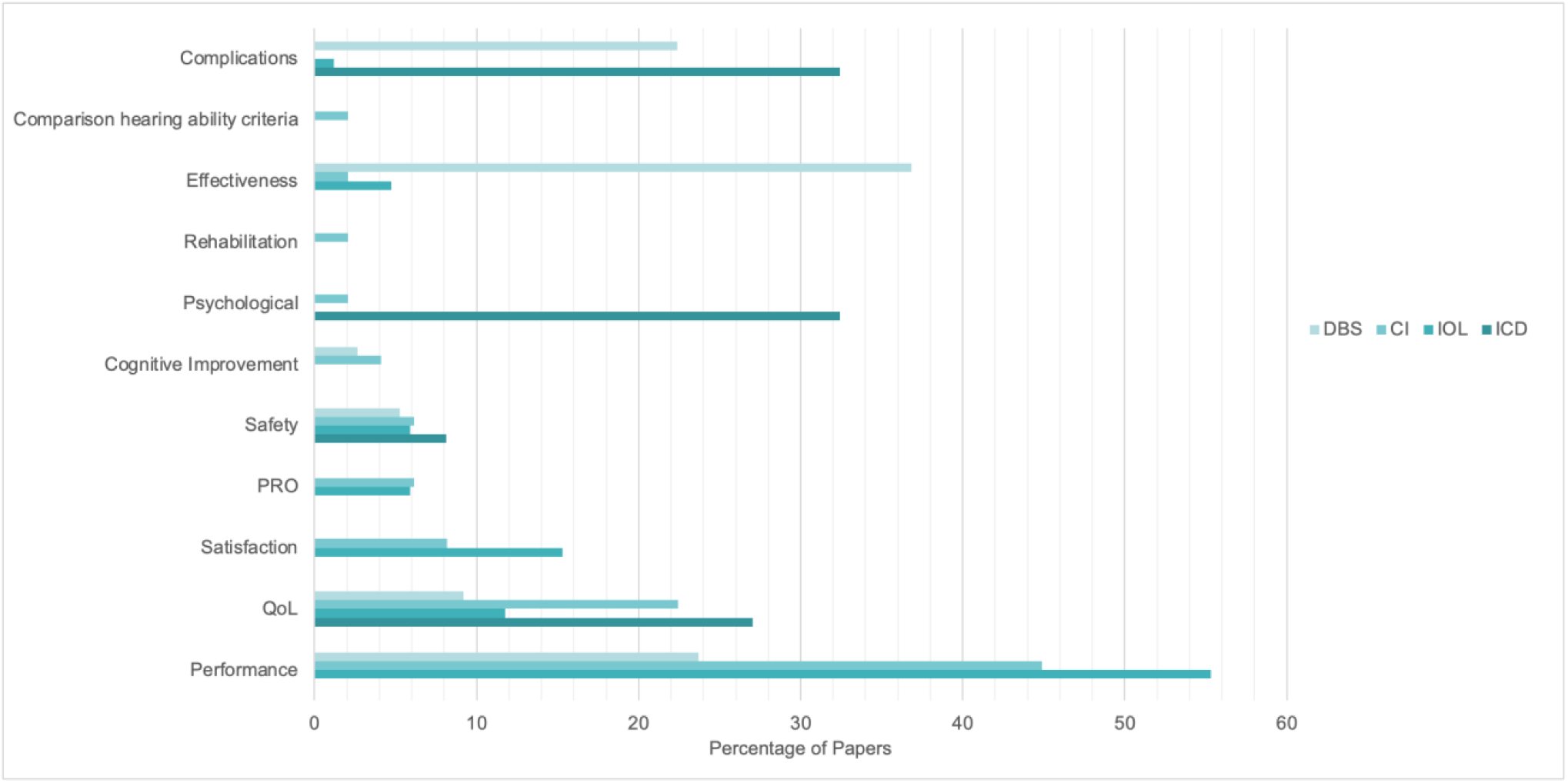
Occurrence of the taxonomy across different implant type studies.

Additionally, we investigated how many studies dealing with either satisfaction, PRO or QoL, also specifically mention the implant manufacturer. This analysis was done for CIs, IOLs and DBS only, as no such data were available for ICDs. The results are depicted in Figure 7. Only 50% of IOL studies dealing with PROs also mention the implant manufacturer with the number decreasing for CIs and DBS. Here only 22% and 14% of papers also mention the implant manufacturer. Despite the ability to perform cross-manufacturer comparisons of PROs, none of those studies reported such comparisons. Considering that all three implant types significantly influence patients’ quality of life this finding is particularly surprising. Clearly, manufacturer comparisons are not straight-forward as they strongly depend on inter-individual parameters which are perhaps difficult to account for. However, we argue that such comparisons can be done when (1) a large number of patients are considered such that inter-individual variability is negligible and (2) attempts to match the patients in terms of condition, age, sex and other influencing factors have been made.

**Figure 7.**
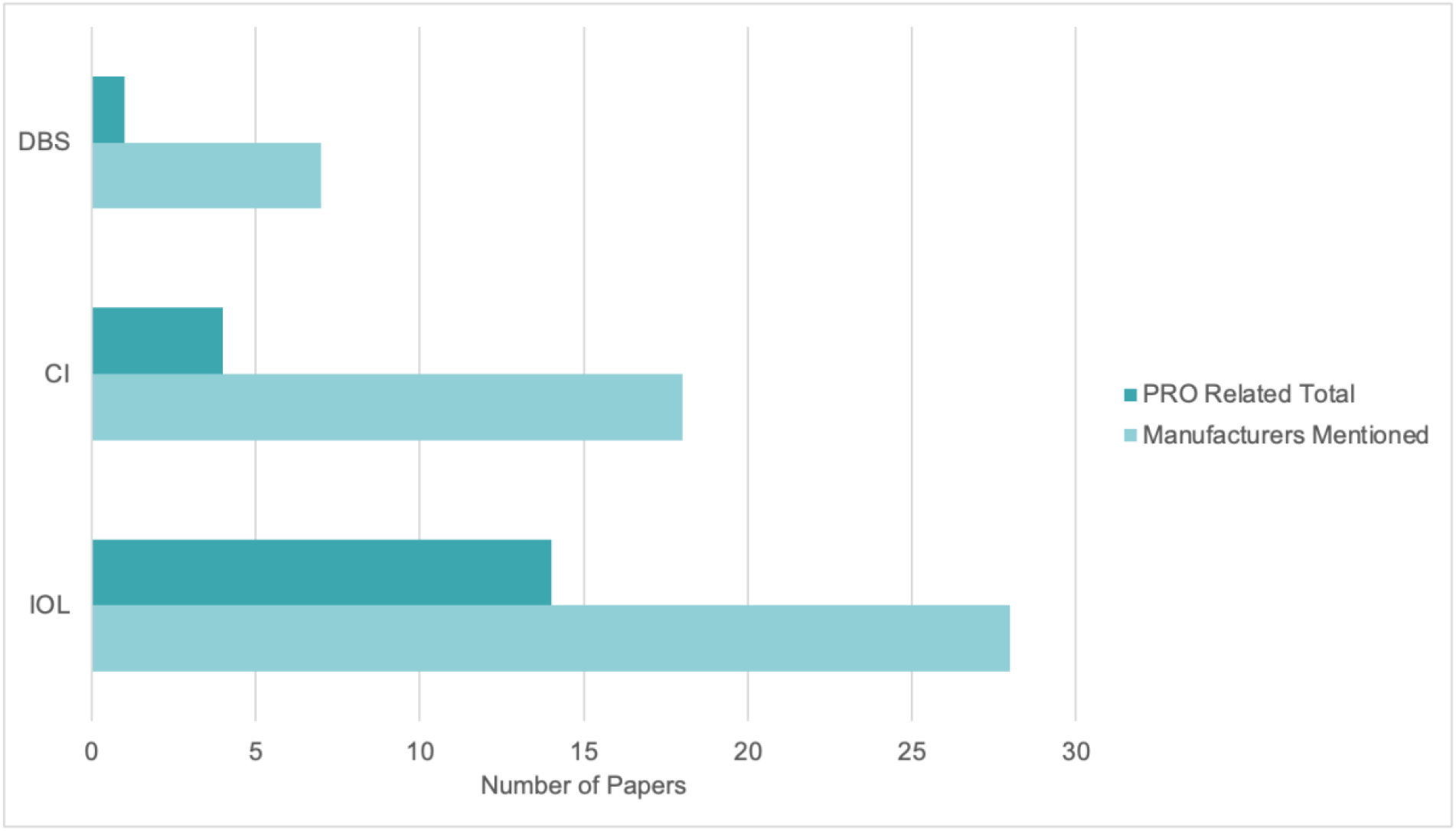
Number of papers mentioning the manufacturers and dealing with some form of PRO.

## Discussion

In this scoping literature review, we found that the majority of papers across implant types were mainly focusing on the performance of the device. The next theme that the studies in this review focused on is QoL assessment. Depending on the implant type, studies focused on different aspects of the implanted devices. Our scoping review also demonstrates that manufacturers of implanted devices were mentioned >95% of the time for DBS and IOLs papers, but only 75% of the time for CIs and <40% of the time for ICDs (see Figure 8).

**Figure 8.**
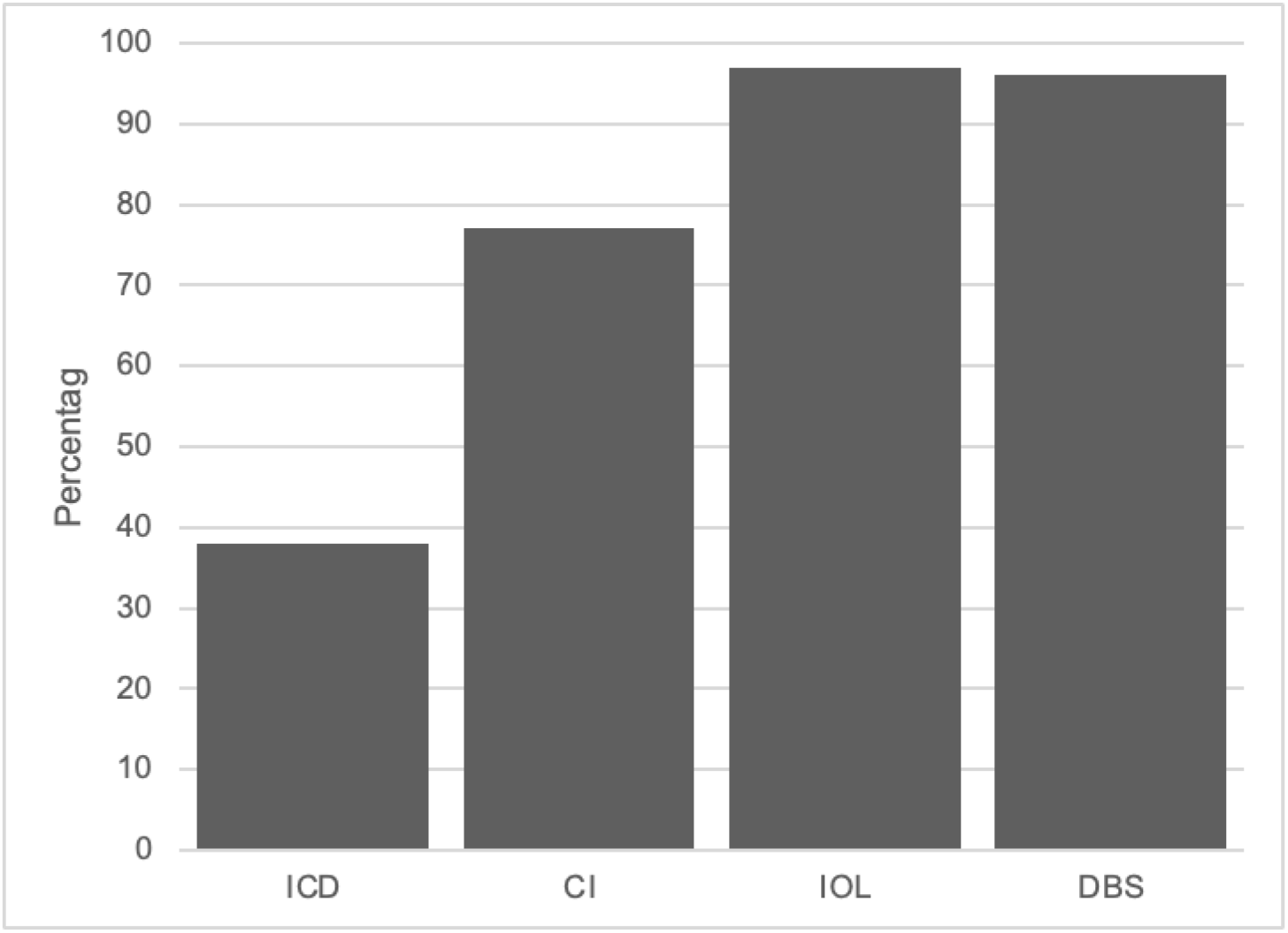
Percentage of reviewed publications mentioning the manufacturer.

However, only 50% of the papers for IOL dealing with PROs and 22% and 14% for CIs and DBS respectively, mentioned the manufacturer in the context of PROs. This is quite surprising as patients, manufacturers and medical professionals alike should be interested in the comparison of implants across manufacturers to be able to make informed decisions when choosing a specific device. This is particularly relevant for implants that directly affect sensory perception, whether or not they stimulate the brain directly. CIs for example, are often implanted in children, thus the specific implant choice can significantly affect patient’s quality of life for many years, as well drive cognitive, emotional, and social development (Humphries et. al., 2012; de Almeida et. al., 2015). Importantly we note that none of the papers we present in this scoping review attempted to perform a direct comparison amongst manufacturers with regards to QoL, despite the fact that data are available, sometimes within one centre, to perform such a comparison. As in all reviews, the selection of search terms strongly influences the spectrum of results achieved. It was critical to choose a very specific search term that could apply for all indications, to enable comparisons of outcomes across these different medical areas. Thus, it is possible that different search terms in each indication specifically would have yielded other publications that perform comparisons between manufacturers. However, a heuristic search we performed for CI, using many different search terms, did not yield any papers that present manufacturer comparisons.

Patients and medical practitioners alike would greatly benefit from easily accessible data and manufacturer comparisons with regards to PROs to make informed decisions prior implantation. Clearly, these comparisons are highly subjective and vary greatly, depending on the individual tested, their medical history as well as the skills of the surgeon and the rehabilitation process following implantation. Yet, this should not prevent the scientific community from attempting to perform such comparisons for the greater benefit of the medical community, patients and physicians alike. Also manufacturers should be more open to allow those types of comparisons as their product would be able to benefit greatly as it allows for constant improvements. Additionally, with sufficient studies and cohort sizes individual differences would eventually cancel out, thus, enabling informed implant choices and recommendations.

Based on the publishing date of the reviewed literature, we may conclude that studies concerning PROs become more relevant especially for CIs, where most of the papers were either published in 2021 or 2020. This testifies for the increased interest of the otolaryngology community in addressing this type of outcome in CI. Similarly, for IOLs, most papers were published in 2017 and 2021, suggesting a similar trend in this field. However, this is not the case for papers on DBS and ICDs. For DBS, most papers are 12 to 16 years old and for ICDs, most papers are 10 to 17 years old. In both fields, the functionality of the sensory system may have a smaller priority in comparison to survival (ICD) and regaining of motor function in movement disorder patients (DBS). The recent advancements in the field of DBS to treat mental health conditions such as Depression and Obsessive Compulsive Disorders (Baldermann et al., 2021; Van Westen et al., 2022; Li et al., 2021; Fenoy et al., 2022; Scangos et al., 2021; Youssef et al., 2021) might drive the field to take a closer look at PROs. Based on the above, we suggest that future research should focus more on PRO based manufacturer comparisons for active as well as passive implants, to allow patients and surgeons to make informed decisions. Further research should also be conducted on the identification of appropriate tests to rate patient satisfaction.

## Conclusions

In this review, we conducted a literature search on the availability of manufacturer comparisons for outcome-based parameters for active and passive implants. This was done as PROs become more important to patients and surgeons when choosing from a selection of implants by different manufacturers. We found that using the search term ‘patient reported outcomes’ led to a variety of results, most of them focusing on the performance of the implant, using objective measurement techniques. Within the papers addressing QoL, PROs and patient satisfaction, less than 50% mentioned the implant manufacturer. Therefore, we suggest that there is a gap in published research for active implants and more research must focus on manufacturer comparisons based on PROs. This is particularly important for medical implants in younger patients as in the CI example since the influence on cognitive, social, and emotional processing has a significant impact on their development. This will lead to an informed decision-making process by patients and surgeons to guarantee the best long-term outcomes. It also fosters innovation as manufacturers are forced to continuously improve their products.

## Data Availability

All relevant data are within the manuscript and its Supporting Information files.

https://docs.google.com/spreadsheets/d/1iQRHgUvgRhrXup1BewfuotDfoCJ8YMNf/edit?usp=sharing&ouid=106617026350517039574&rtpof=true&sd=true

## Acknowledgments

The authors state no conflict of interest. Syte receives financial compensation for consultation to Cochlear, MedEl and Abbott in the past.

